# Utility of 3D Facial Analysis As A Biomarker In Rare Diseases Exploration with Hereditary Angioedema

**DOI:** 10.64898/2026.03.26.26349135

**Authors:** Saumya Jamuar, Richard Palmer, Lee Haur Yueh, Faith Li-Ann Chia, Choo Beng Goh, Stuart Lee, Petra Helmholz, Shermaine Chan, Gareth Baynam

## Abstract

**Importance:** Digital Phenotyping (DP) utilizes digital technologies to assess observable phenotypic traits, enhancing our understanding of various disease states. It aims to support equitable interventions through diverse digital data, fostering inclusivity by integrating various digital devices. Central to DP is identifying digital biomarkers (DBMs), which offer real-time health monitoring and personalized insights into disease conditions. Applying machine learning (ML) techniques on non-invasive signals sets the ideal platform for precision medicine application, especially in rare diseases (RDs).

People living with rare diseases (PLWRD) often face significant challenges in receiving timely and accurate diagnoses, leading to what is known as a diagnostic odyssey. Digital phenotyping (DP) offers a promising solution by leveraging advanced technology, such as 3D facial photography, to capture unique digital signatures associated with various rare diseases. This innovative approach not only aids in the identification of these conditions but also facilitates the detection of digital biomarkers (DBM). These biomarkers enable healthcare providers to monitor the progression of the disease over time, enhancing patient care and potentially shortening the duration of the diagnostic odyssey. By utilizing DP and DBMs, we can improve both the diagnosis and management of RDs, ultimately leading to better health outcomes for affected participants.

**Objective:** To identify whether DBMs can be identified by DP utilizing 3D facial imaging techniques in outpatient settings in participants with RDs.

The primary objective of this study was to determine if specific facial measurements in participants with RD who experience transient episodes of facial swelling (oedema) differ from established ethnically matched norms. The secondary objective was to assess peri-orbital and/or facial swelling as a potential biomarker for identifying flare-ups in hereditary angioedema (HAE).

**Design, setting, and participants:** This multicentre observational study was conducted in 3 hospitals in Singapore. The eligible participants were male and female RD participants of various age groups. The study duration was 4 years and 8 months.

Interventions: Twenty participants of Chinese genetic ancestry were photographed using a 3D camera. Additionally, two participants with hereditary angioedema (HAE) were photographed during acute stages of disease flare-ups.

**Main outcomes and measures:** The obtained facial scans of participants (that included participants with HAE in non-acute phase) were plotted using Artificial Intelligence-powered software - Cliniface. The growth curves and facial landmarks obtained were compared against the growth curves of normal RD-unaffected individuals of Chinese genetic ancestry. The two participants with HAE were photographed qualitatively over a longer period of time, and their scans were plotted, yielding growth curves.

**Results:** Distinct facial markers such as periorbital swelling were identified in two qualitatively assessed HAE participants during flare-up stages. This provides an opportunity to explore and validate further if these facial signatures in a disease condition can be assigned as DBM for HAE.

**Conclusions and relevance:** This study explores the utility of 3D facial analysis as a DBM in rare diseases such as HAE. Applying non-invasive signals coupled with AI may open new vistas for precision medicine in real-world settings.

The individual measurements that yielded small p-values demonstrate significant relevance and potential utility. These findings offer preliminary objective evidence that supports existing subjective reports of facial features in the literature. Additionally, while DP’s diagnostic capabilities may be limited, it successfully identified DBM, which could facilitate disease monitoring in conditions such as HAE.

**Author Summary:** Rare diseases pose a significant challenge to all stakeholders, including clinicians, patients’ families, care providers, and the healthcare system. Diagnostic delays are integral and impose a massive financial and emotional burden on everyone involved in care delivery, beyond the patients themselves. A universally acceptable, scalable, and replicable non-invasive mechanism to detect distinct biomarkers associated with a rare disease can help identify the disease’s signs and symptoms far earlier and ease the burden. An easy-to-deploy approach could be 3D facial imaging of patients with rare diseases, which are associated with distinct or subtle facial changes at different stages of disease progression. A rare disease, such as hereditary angioedema, which is known to exhibit facial swelling in patients during the acute disease state, is a prime example. The facial changes can be identified and assigned as specific disease markers, also known as facial biomarkers. These facial biomarkers can be identified and measured using 3D facial imaging when patients present to the clinic. Subsequently, these initial signs can be correlated clinically to establish a firm diagnosis earlier than traditional approaches.

## Introduction

People living with rare diseases (PLWRDs) often undergo a prolonged diagnostic odyssey, often lasting more than 4 years from the initial presentation, despite advances in genomic testing capabilities [1]. Part of the reason is the sheer number of rare and ultra rare diseases (RDs), estimated to be >10,000 [2], which make it impossible for any clinician to be familiar with all of them. In addition, many of the phenotypic features of these rare diseases overlap with other commoner diseases, often leading to misdiagnosis [3].

Advancements in technologies such as digital phenotyping (DP) using 2D and 3D photogrammetry have been used to identify facial signatures associated with these rare disorders, which in turn helps to reduce this diagnostic odyssey [4]. In addition, DP provides an opportunity to identify digital biomarkers (DBM), which can then be used to objectively monitor the progression of a disease and/or response to treatment, both in the clinical trial as well as real world setting.

Hereditary angioedema (HAE) is a rare genetic disease characterised by intermittent swelling of the soft tissues due to a defect in the SERPING1 protein. The swelling typically involves the periorbital and perioral areas. However, if the swelling involves the airways, it can be life threatening and requires immediate medical care. However, patients with HAE often go undiagnosed, and even when diagnosed, due to lack of objective biomarkers to define exacerbation often receive delayed treatment, not only leading to increased anxiety among participants, but also putting them at risk of life threatening complications. In this study, we aimed to understand the role of 3D photography in diagnosis and monitoring of participants with HAE.

RDs are compounded by high mortality rates and significant disability, leading to substantial societal costs [5]. Due to the limited number of subjects eligible for clinical studies, typically small, single-arm, non-randomized, and open-label approaches are undertaken [6]. Also, the outcome measures are often subjective and can depend on both patient motivation and the rater’s assessment [7]. Furthermore, outcomes typically represent the patient’s condition at a specific point in time, whereas the severity of symptoms can fluctuate over time [8]. The challenge of clinical heterogeneity significantly hampers the establishment of reliable clinical endpoints in rare disease trials. Small and fragmented patient populations often present insufficient data, making it challenging to define trustworthy outcome measures. Additionally, the constraints of limited diagnostic tools, challenges in identifying eligible participants, and variability in disease progression collectively lower enrolment rates. This leads to extended study timelines and compromises data integrity, ultimately contributing to a higher failure rate in RD trials compared with trials for more common conditions [9,10]. To improve the success of clinical research in this underserved area, the precision and objectivity of outcome measurements are crucial. Digital BioMarkers (DBM) could address this unmet need to some extent.

Establishing DBM in rare diseases (RD) using non-invasive methods, such as 3D facial imaging, is a novel emerging concept. In this study, we present our exploratory work of an earlier hypothesis concerning the utility of 3D facial analysis as a digital biomarker [4] in those with Chinese, Malay, and Indian genetic ancestry. The primary objective of this study was to determine whether facial measurements in PLWRD, characterized by transient episodes of facial oedema, deviate from ethnically matched established norms. Additionally, we sought to evaluate peri-orbital swelling as a potential biomarker for identifying HAE flare-ups.

## Results

### Demographics

A total of 7 participants with HAE were recruited and photographed during the study period. Out of these, 2 participants had exacerbations and serial photographs were obtained for both participants. There were 20 participants of Chinese, 1 of Malay and 1 Indian genetic ancestries.

In addition, we recruited 15 participants of Chinese genetic ancestry with other similar RDs (**Table 1**).

**Table 1.** Recruited participants of Chinese genetic ancestry with RDs other than hereditary angioedema.

| Participant | Age category | Sex | Diagnosis |
| --- | --- | --- | --- |
| 1. | Paediatric | Male | Mucopolysaccharidosis type II |
| 2. | Paediatric | Male | Achondroplasia |
| 3. | Adult | Female | Incontinentia pigmenti |
| 4. | Adult | Female | Incontinentia pigmenti |
| 5. | Paediatric | Male | Mucopolysaccharidosis Type VI |
| 6. | Paediatric | Male | Mucopolysaccharidosis Type VI |
| 7. | Paediatric | Male | Mucopolysaccharidosis Type VI |
| 8. | Paediatric | Female | Mucopolysaccharidosis Type IV - Morquio A syndrome |
| 9. | Paediatric | Female | Achondroplasia |
| 10. | Paediatric | Female | Velocardiofacial syndrome |
| 11. | Paediatric | Male | Achondroplasia |
| 12. | Paediatric | Male | Noonan syndrome |
| 13. | Paediatric | Male | Noonan Syndrome |
| 14. | Paediatric | Female | Achondroplasia |
| 15. | Paediatric | Male | Noonan Syndrome |

### Use of 3D facial analysis for diagnostics in HAE and other rare diseases

The test cohort comprised 20 participants of Chinese genetic ancestry (five participants of Chinese ancestry with HAE and 15 with other rare diseases as shown in **Table 1**). The test cohort exhibited wide variation in the degree of facial oedema (or similar features) present. Due to the relatively small test sample size, it was not possible to test for sex differences, and all analyses were performed against measurement growth curves regressed against the combined male and female Chinese genetic ancestry reference dataset.

The objective analysis component of our study with quantitative exploratory comparisons had 108 measurement variables analysed from 58 base measurements. Of the 108, 33 variables were rejected for failing the normality test. P-values were calculated for the remaining 75 variables and of these, 11 had p-values lower than the significance threshold of 0.05. After FWER correction over the 75 variables, a single one – the Outer Canthal, Nasal Angle retained a significant p-value of 0.01 allowing for the null hypothesis of equivalency with the control cohort to be rejected. The 11 most significant results (with Holm-Bonferroni unadjusted p-values lower than 0.05) are presented in **Table 2**. The complete table of results is provided in the supplementary material. Mean effect sizes are given in terms of the standardised (Z) scores with the margin of error (ME) calculated at 95%.

**Table 2.**
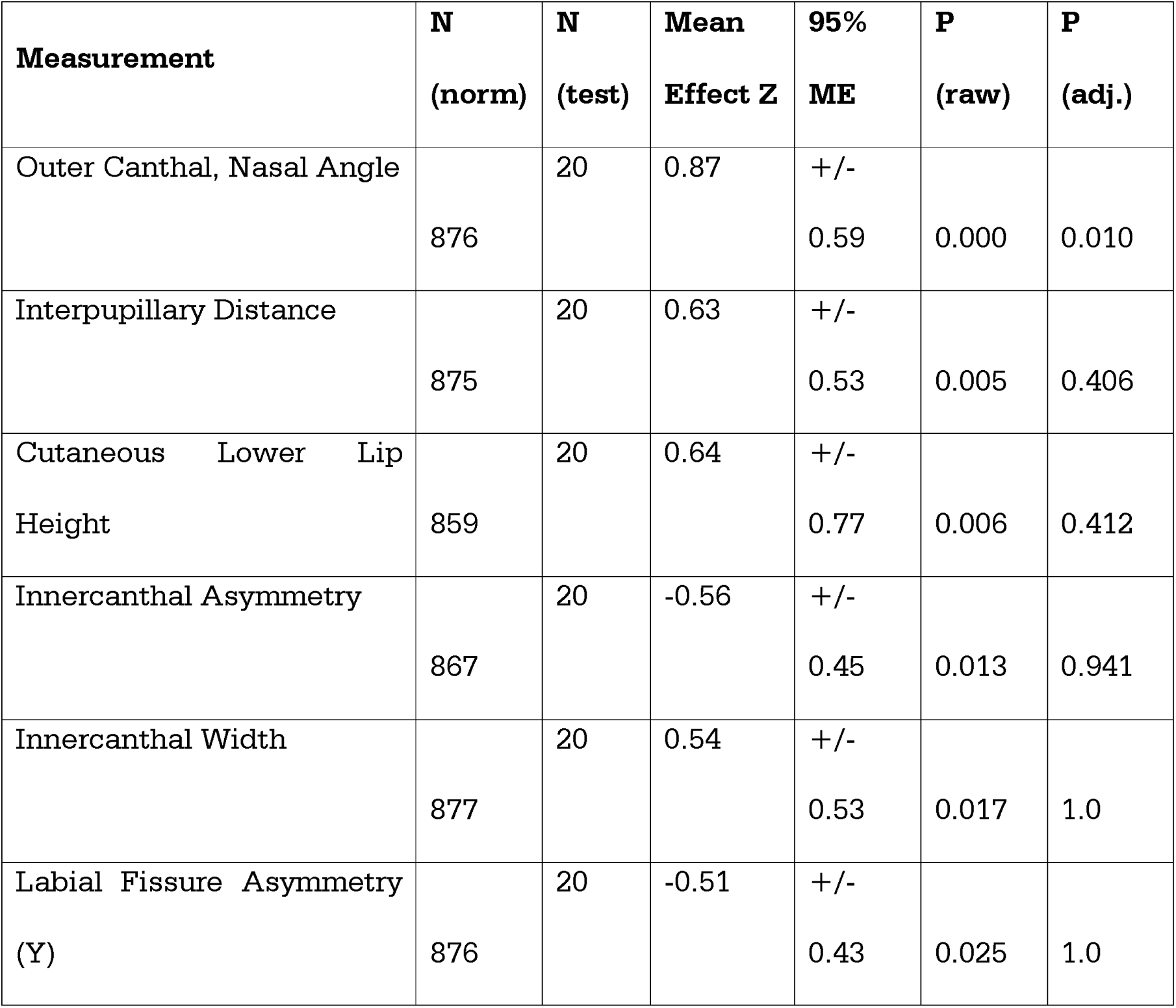
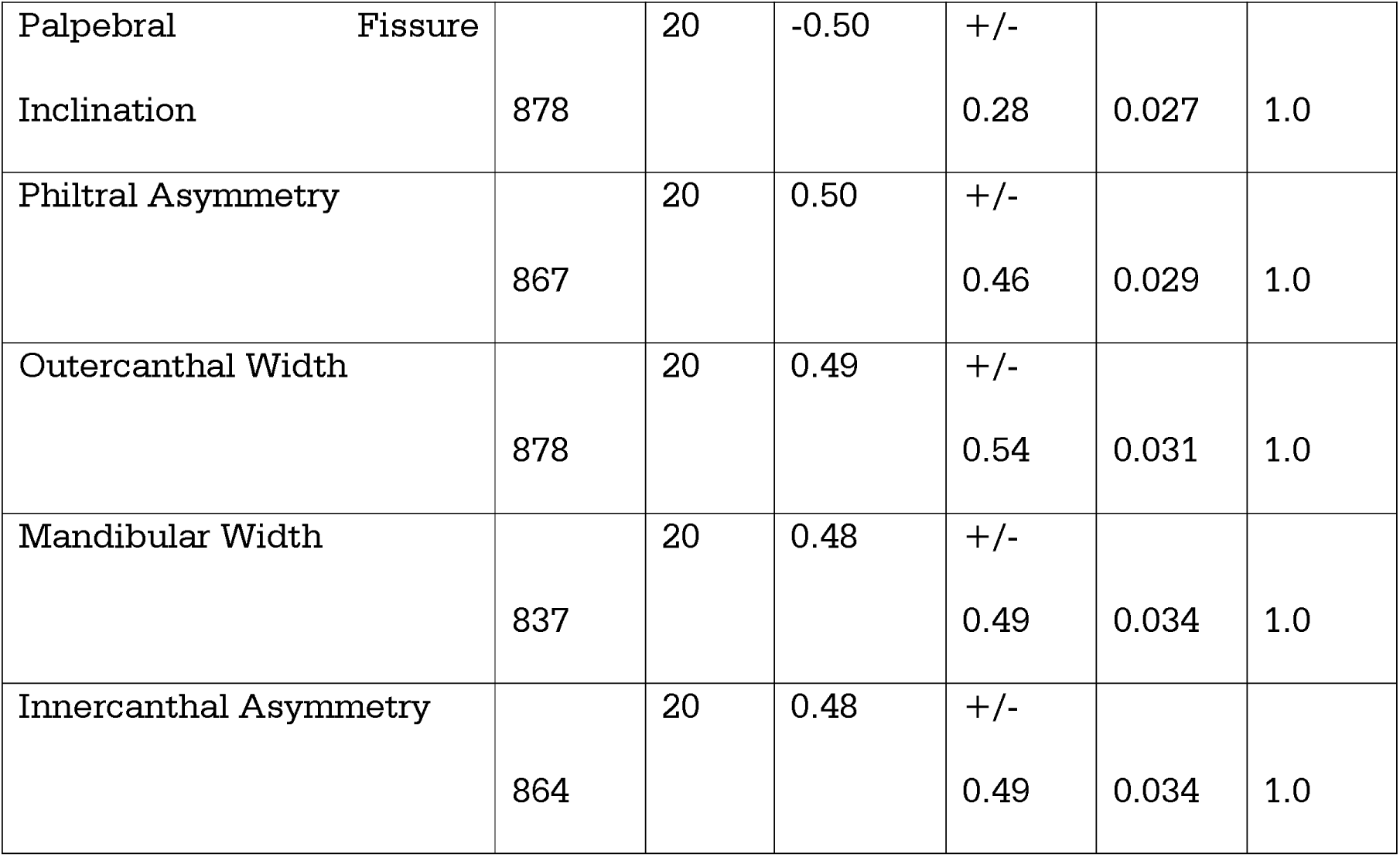
Statistically significant variables and their values.

The measurements of the Outer Canthal, Nasal Angle from both the control and test cohorts (test observations as black stars) are plotted in **Figure 1**. Especially in the case of the younger participants in the test cohort, the observed values are generally larger than the age matched regressed mean. While this result is statistically significant, due to the small sample size and variation in the character of the test cohort further investigation using independently collected data is necessary to corroborate this outcome.

**Figure 1.**
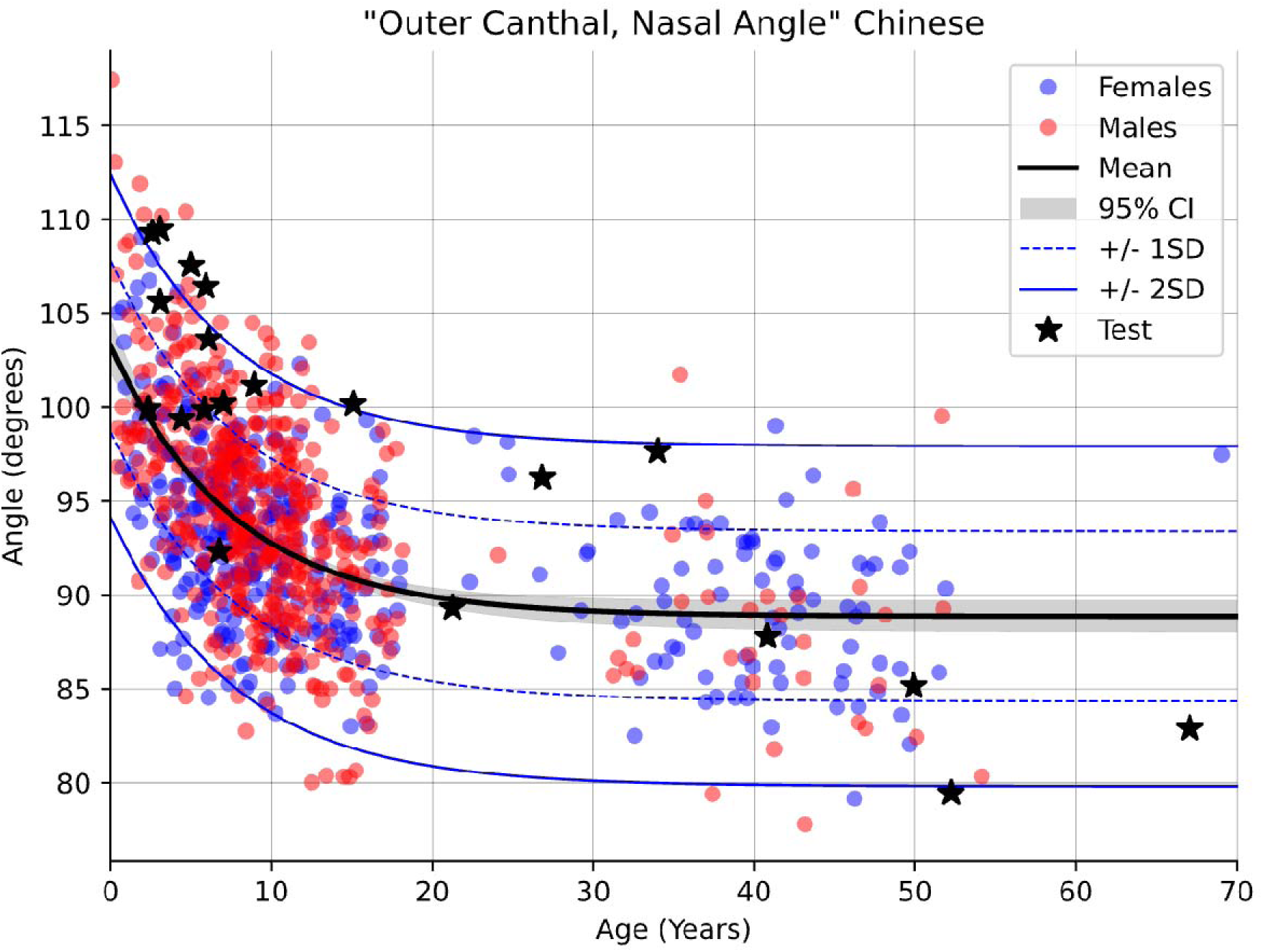
Measurement of the Outer Canthal, Nasal Angle in both the control and test cohorts (test observations denoted by black stars).

Of the 75 admissible measurement variables analysed, only the Outer Canthal, Nasal Angle showed a statistically significant difference (after FWER correction) between healthy participants of Chinese genetic ancestry, and those with HAE or other closely mimicking genetic conditions. This suggests that 3D facial imaging may be useful in helping to identify participants who experience facial swelling during remission. Across all other measurements analysed, the observed differences were too limited to yield clear, statistically significant results. Our sample size was small due to the limited population in Singapore and the restricted number of participants with RDs. Apart from the Outer Canthal, Nasal Angle, the results in **Table 2** suggest that at least ten other measurements (having small p-values) - including measurements of specific facial asymmetry - may help to objectively indicate the presence of facial oedema (or facial traits presenting similarly). These results suggest the potential for measurements derived from 3D facial images to monitor resolution of facial oedema in remission and/or detect facial oedema during mild/subclinical exacerbations. Independent follow-up investigations of these measurements will likely help to corroborate these findings.

### Use of 3D facial analysis for monitoring in HAE

A long-term qualitative assessment of serial images of participants with HAE was conducted to identify distinct facial markers associated with the disease. These two different, but complementary, approaches were necessitated by the scarcity of rare disease cases and the practical complications of capturing facial images of HAE participants during severe and infrequent episodes of acute facial oedema. 3D facial images were captured from two participants over a longer duration to be able to identify markers of facial appearance change likely associated with HAE.

### Participant 1

‘A male, adult participant of Malay genetic ancestry, was photographed at seven time points over 687 days, including images taken immediately after a facial oedema flare (**Table 3**). Images were imported into Cliniface for Procrustes analysis and visualisation of shape change over time.

**Table 3.** The Imaging timelines of the male, adult participant with HAE during qualitative assessment.

| Event | Timeline |
| --- | --- |
| Baseline 1 | Day 1 |
| Baseline 2 | Day 154 |
| HAE flare started @ 18:00 – admitted<br>@ 22:00 | Day 427 |
| Post 1 | Day 428 |
| Post 2 | Day 429 |
| Baseline 3 | Day 589 (from HAE flare day 1) |
| Baseline 4 | Day 687 |

\*baseline = participant did not complain of exacerbation

In our analysis of 3D facial images from a male participants with HAE (**Figure 2**), we initially identified minimal differences in facial volume between the first two baseline measurements taken five months apart (**Figure 3).** A significant observation was noted during the transition from baseline measurements to the first post-flare assessment: asymmetric peri-orbital swelling, with an increase of more than 3 mm around the right eye. This swelling around the right eye showed some reduction one day later in the second post-flare assessment, while the swelling around the left eye remained unchanged, aside from a slight reduction observed in the left cheek. At the third baseline measurement, taken six months after the flare, the swelling around the right eye had significantly decreased, with a change greater than 3 mm from the second post-flare assessment. There was also a slight increase in swelling around the left eye, which appeared to resolve during the fourth baseline measurement taken three months later (it was later noted that the participant was non-compliant with medication at baseline three, and might have had a mild exacerbation which did not warrant any medical attention). When comparing the second baseline measurement taken 14 months earlier, we found that the periorbital swelling had not completely subsided, although the asymmetry was no longer present. Given the time elapsed since the previous measurements, the findings at baseline four fell within the expected range of variation. Overall, these observations suggest that 3D facial analysis is effective in differentiating between flare and non-flare states, as well as in assessing treatment responses, particularly in instances of non-compliance. Additionally, we noted slight symmetric changes around the cheekbones and lower eyelids, along with a retrusion in the labial fissure, which may have been influenced by the participant’s facial expression (slight smile). The final image, captured one day after the second exacerbation, exhibited a similarity in peri-orbital swelling to that seen after the first exacerbation. However, an increase in swelling was observed in the cheeks. The apparent sublabial retrusion is likely an artifact resulting from the average mask fitting being positioned more anteriorly, attributed to the increased extrusion around the eyes and cheeks.

**Figure 2.** 3 D facial images of adult male participant with HAE during different stages of diseases over a span of 687 days.

**Figure 3.**
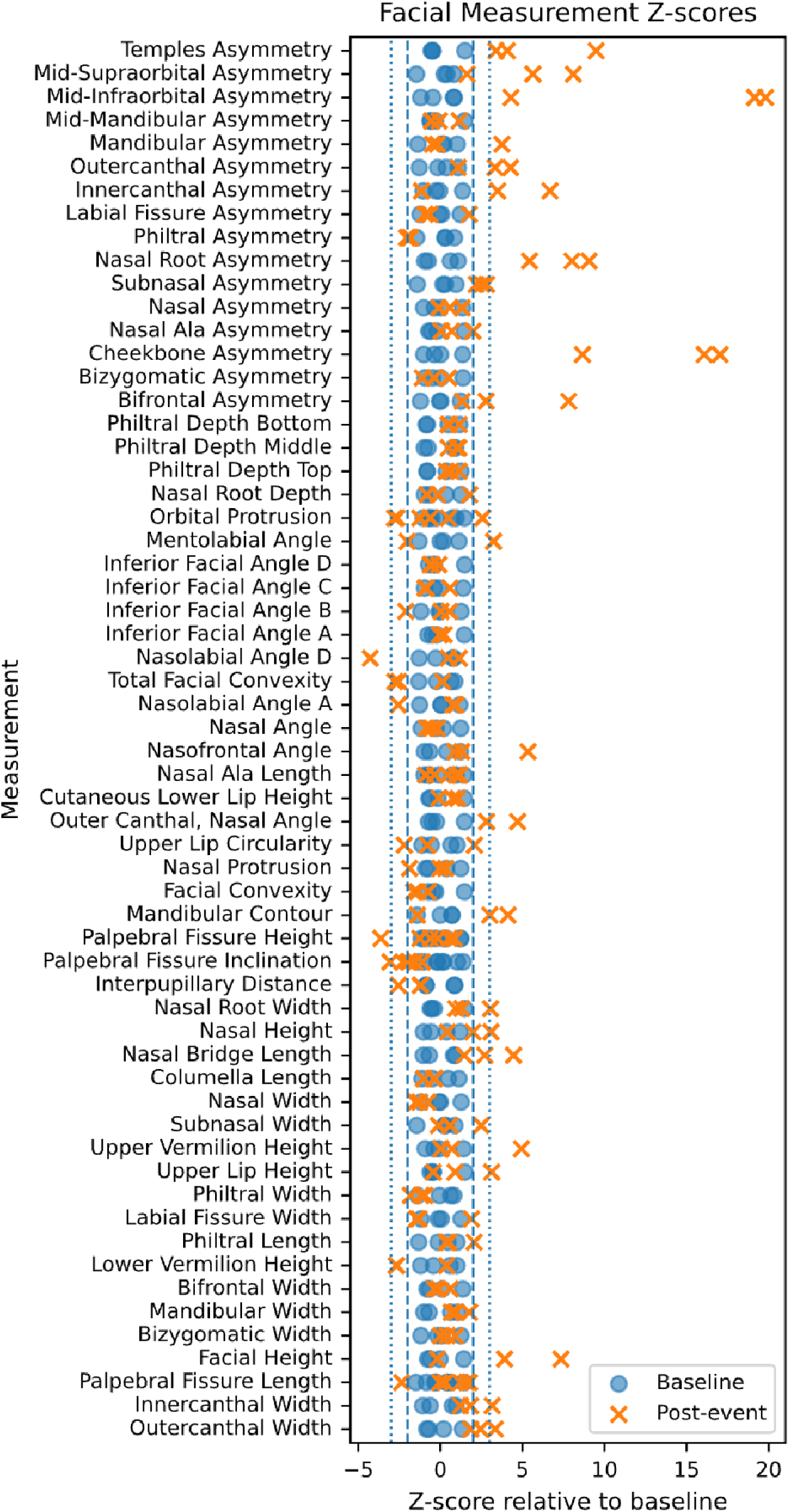
Comparison of facial landmarks and Z-score deviation relative to mean baseline.

Differences between the participant at baseline (4 images) and after experiencing an exacerbation are summarised in Figure 3. Variation in each measurement across the seven datapoints is shown with respect to the mean of the four baseline datapoints.

### Participant 2

The second participant was female, adult, of Indian genetic ancestry. She was photographed twice, 261 days apart. The first image was taken immediately after she experienced an acute episode of HAE oedema. A baseline image was taken 261 days later for comparison. The images were imported into Cliniface for procrustes analysis and visualisation of facial shape change between these two timepoints.

The second participant with HAE facial imaging analysis revealed asymmetric peri-orbital swelling (greater on the participant’s left). Asymmetric swelling of cheeks and lower jaw – were also more pronounced on participant’s left side. Apparent retrusion in midline, and apparent compensatory retrusion in the participant’s right upper eyelid seems likely to be an artefact of the average mask fitting (**Figure 4**). The facial measurement z-scores could not be determined for the second participant as there were only two sets of images for comparison. The asymmetry findings, suggest that approaches that rely only on landmarks, such as crow’s feet landmarks, and do not assess symmetry may miss critical indicators of disease flare/ treatment response/ compliance.

**Figure 4.** 3 D facial images of a female participant with HAE at an interval of 261 days.

## Discussion

Our study conducted an exploratory analysis of facial morphology in participants with RDs, specifically focusing on participants of Chinese genetic ancestry. A total of 20 participants with various genetic conditions associated with facial oedema were compared to a demographically matched control cohort using 3D facial photogrammetry. Facial images were analysed for 58 base measurements, generating 108 measurement variables, demonstrating opportunities to explore, identify, and differentiate syndrome-specific apparent and subtle changes in microdetails. These changes need to be established for their distinct and identifiable signatures during evolutionary or resolutory phases of different RDs and in common diseases. The supplementary table enumerates all the tested variables which shows that our study was rigorous and we took into consideration only robust indicators from participant groups. Wider-scale studies need to be conducted on healthy individuals and RD participants hailing from different genetic ancestries, genders, and age groups for comparisons and harmonizations of clinically significant changes associated as identifiable signatures for specific syndromes.

Our study’s results demonstrate findings in symmetric and asymmetric facial features that correlate with disease flare, resolution and treatment non-compliance, and that overlap between the imaged participants. This suggests that 3D facial analysis using the Cliniface software is useful in discriminating HAE flare (acute episode) from non-flare states. The nature of the swelling shown in the post-flare images in both individuals is similar. Both participants have notable asymmetric peri-orbital swelling (though on different sides), and swelling of the cheeks and lower jaw is also apparent in both. From these observations we infer that periorbital swelling and asymmetry during acute episode of HAE through 3D facial imaging could be a potential DBM to predict the course of the disease.

While promising, the results of this study are limited by the small sample size and inherent variability in the test cohort due to incorporation of different disease types. Larger studies with diverse participants is important to demonstrate applicability of these results across different populations. However, given that we used the participants as their control, DBM identified through this study are biologically plausible and can potentially be used for DP in HAE.

DP and DBM identified through 3D facial analysis, hold significant promise for disease screening, early disease detection, monitoring, and personalized treatment. Our study provides an exploratory utility of 3D facial imaging as a DBM for assessing disease states, treatment response, and patient compliance, particularly in RDs. We have shown preliminary and tentative evidence (quantitative and qualitative) for efficacy of DBM from 3D facial images for the objective detection of latent facial oedema. While challenges such as small sample sizes and limited genetic ancestry diversity persist, ongoing data collection and technological advancements require further broad-based investigations, validation, and applications. Cliniface’s precision underscores the potential of digital tools in advancing precision medicine and avenues for improving patient outcomes.

## Materials and Methods

This was a prospective cohort study with two different components. The first component is a quantitative case-control analysis, and the second is a prospective qualitative study.

### Participant recruitment

Participants were recruited across three hospitals in Singapore, KK Women’s and Children’s Hospital (KKH), Singapore General Hospital (SGH), and Tan Tock Seng Hospital (TTSH), under an IRB-approved study (SingHealth CIRB 2021/2419). Participants (or their legal guardians if the participant was under 21 years) consented to participate in the study. The study duration was 4 years and 8 months.

### Cohort selection

Participants with clinical and/or genetic diagnosis of HAE were recruited for the study. As HAE is a RD, we expanded our cohort by recruiting participants with other RDs that had similar clinical phenotypic features such as facial swelling and/or facial asymmetry. This included participants with mucopolysaccharidosis, Noonan syndrome, incontinentia pigmenti, velocardiofacial syndrome and achondroplasia.

### Digital phenotyping

A 3D facial image of each participant was captured by a member of the study team in Singapore using Vectra M3 (https://www.canfieldsci.com/imaging-systems/vectra-m3-3d-imaging-system/) camera system. These images were analyzed with the Cliniface software program to extract facial landmarks and 58 different measurements [11]. The resulting de-identified data were transmitted to the Cliniface team in Western Australia (WA). All participants with HAE were photographed at baseline, and then were photographed serially during exacerbation(s), wherever possible.

The method involved an exploratory analysis of variation across 58 different facial measurements of a disease-affected cohort versus typical (unaffected) reference values derived from regression modelling of the demographically matched (age/sex / genetic ancestry) population. For each measurement variable (some measurements being comprised of several variables corresponding to different spatial dimensions), two-tailed t-testing of the test (RD affected) cohort was performed against the regressed model of the corresponding measurement variable derived from a control (RD unaffected) cohort. A total of 108 different measurement variables were investigated. Data for the control cohort were generated as part of a separate study [12] and were used to regress growth curves for each of the measurement variables against which the test cohort could be compared. The growth curve regression methodology followed that described in Jamuar et al [12].

Statistical analysis of variation between the test and control cohorts used two-tailed t-testing since it was not possible to state the expected direction of variation in each measurement from its norm with certainty beforehand.

For each measurement variable, the analysis proceeded as follows:

1. The control cohort was regressed to estimate the population mean and standard deviation (SD) across the full age range (0-70 years). Outliers exceeding 3 SD were removed, and regression was then performed again without them to produce a growth curve.
2. A z-score for each observation (participant) in the test cohort was calculated by comparing to the regressed growth curve at the corresponding age.
3. The test cohort z-scores were assessed for normality (skewness and kurtosis), and the test was rejected if the z-scores were not normally distributed (P < 0.05).
4. The null-hypothesis for the test cohort z-scores was defined as being zero (i.e., equivalent to the regressed mean of the control cohort).
5. A two-tailed t-test was performed with absolute values of the t statistic greater than the critical value of 2.093 (DF =19), denoting rejection of the null hypothesis (a significance level of 0.05). The corresponding p-value was found from the cumulative distribution function of the t-distribution.

After deriving p-values for all measurements that were not rejected out of hand by normality testing, the complete set was adjusted using the Holm-Bonferroni method to control the family-wise error rate (FWER). The Holm-Bonferroni method was used as it is conservative and does not require the set of variables to be independent from one another (correlations between variables being likely due to different measurements being extracted from very nearby regions of the face across the same set of individuals). All statistical analyses were performed using Python (version 3.12) supported by the SciPy (version 1.16.3) and stats models (version 0.14.5) libraries.

## Limitations

Our study has several shortcomings. Only 20 participants in our comparative study were of Chinese genetic ancestry, and of these only five of these individuals had HAE. A large and diverse pool of participants and comparison across different ethnic groups is crucial for increasing confidence in the results obtained. In a Chinese genetic ancestry majority country such as Singapore, with a small population base, this was inevitable. The normal subject comparison arm comprised mainly paediatric subjects, while the diseased state participant group included some adults also. Our qualitative analysis of participants with HAE was limited to two participants only. Whilst the HAE participant numbers were small, the use of multiple images per affected participant and the consistency of findings between the affected participants (swelling, asymmetry) are supportive and biologically plausible. We hope to further establish our experimental findings through ongoing data collection from participants of different genetic ancestries and disease conditions to identify additional DBMs.

## Conclusion

DP and DBM identified through 3D facial analysis, provides possible avenues for disease screening, early disease detection, monitoring, and personalized treatment through non-invasive means. Our study provides an exploratory utility of 3D facial imaging as a DBM for assessing disease states, treatment response, and patient compliance, particularly in RDs. We have exhibited preliminary and tentative evidence (quantitative and qualitative) for efficacy of DBM from 3D facial images for objective detection of latent facial oedema.

While challenges such as small sample sizes and limited genetic ancestry diversity persist, ongoing data collection and technological advancements require further broad-based investigations, validation, and applications. Cliniface’s precision underscores the potential of digital tools in advancing precision medicine and improve patient outcomes.

## Data availability

No. Participant-level data on which the analysis was based will not be shared to protect participant anonymity.

## Patient consent

Patient consent was obtained beforehand and permission to use the facial images was obtained in writing on institutional consent form.

## Acknowledgments

The authors thank all the study subjects for participating and cooperating in conducting this study.

Charis Lim, Jade Deng, Melissa Wong, Krishna Kahol Carbajo, and Mia Comerford assisted with the extraction of photographs.

Dr Suchitra Kataria from Melange Communications Pte Ltd, Singapore, provided medical writing support for the manuscript funded by Takeda Pharmaceuticals (Asia Pacific) Pte Ltd.

## Author contributions

All the authors contributed equally to the manuscript writing, editing, and proofreading.

---

**Supplementary table.** Results for all tested variables (rejected samples not shown).

| Measurement | Dimension | N<br>(norm) | N (test) | Effect Z | +/- 95%<br>ME | P (raw) | P<br>(adj.) |
| --- | --- | --- | --- | --- | --- | --- | --- |
| Outercanthal Width | 0 | 878 | 20 | 0.4907 | 0.5373 | 0.0308 | 1 |
| Innercanthal Width | 0 | 877 | 20 | 0.5407 | 0.5279 | 0.0173 | 1 |
| Palpebral Fissure Length | 0 | 878 | 20 | 0.1712 | 0.5708 | 0.4517 | 1 |
| Facial Height | 0 | 859 | 20 | 0.2282 | 0.4976 | 0.3138 | 1 |
| Bizygomatic Width | 0 | 852 | 19 | -0.0807 | 0.5676 | 0.7292 | 1 |
| Mandibular Width | 0 | 837 | 20 | 0.4816 | 0.488 | 0.0336 | 1 |
| Philtral Length | 0 | 875 | 20 | 0.369 | 0.5286 | 0.104 | 1 |
| Labial Fissure Width | 0 | 878 | 20 | -0.2708 | 0.4206 | 0.2299 | 1 |
| Philtral Width | 0 | 875 | 20 | -0.2767 | 0.4022 | 0.2195 | 1 |
| Nasal Width | 0 | 878 | 20 | 0.1002 | 0.4756 | 0.6576 | 1 |
| Columella Length | 0 | 876 | 20 | -0.2105 | 0.5208 | 0.3531 | 1 |
| Nasal Bridge Length | 0 | 877 | 20 | -0.3908 | 0.5235 | 0.085 | 1 |
| Nasal Height | 0 | 877 | 20 | -0.1139 | 0.5042 | 0.6148 | 1 |
| Nasal Root Width | 0 | 875 | 20 | 0.2865 | 0.4928 | 0.206 | 1 |
| Interpupillary Distance | 0 | 875 | 20 | 0.6312 | 0.5252 | 0.0055 | 0.4064 |
| Palpebral Fissure Inclination | 0 | 878 | 20 | -0.4967 | 0.2774 | 0.0269 | 1 |
| Palpebral Fissure Height | 0 | 875 | 20 | -0.2751 | 0.5595 | 0.2263 | 1 |
| Mandibular Contour | 0 | 833 | 20 | -0.3171 | 0.5153 | 0.1622 | 1 |
| Facial Convexity | 0 | 848 | 20 | -0.0292 | 0.5053 | 0.8976 | 1 |
| Nasal Protrusion | 0 | 875 | 20 | 0.145 | 0.5583 | 0.5235 | 1 |
| Upper Lip Circularity | 0 | 864 | 20 | -0.0355 | 0.48 | 0.8749 | 1 |
| Outer Canthal, Nasal Angle | 0 | 876 | 20 | 0.8722 | 0.5869 | 0.0001 | 0.0102 |
| Cutaneous Lower Lip Height | 0 | 859 | 20 | 0.6408 | 0.7703 | 0.0056 | 0.4124 |
| Nasal Ala Length | 0 | 874 | 20 | -0.0625 | 0.5027 | 0.7826 | 1 |
| Nasofrontal Angle | 0 | 864 | 20 | -0.1492 | 0.3825 | 0.5081 | 1 |
| Nasal Angle | 0 | 875 | 20 | 0.3645 | 0.4489 | 0.1067 | 1 |
| Nasolabial Angle A | 0 | 877 | 20 | 0.1583 | 0.5103 | 0.4854 | 1 |
| Total Facial Convexity | 0 | 847 | 20 | 0.3183 | 0.5486 | 0.1619 | 1 |
| Inferior Facial Angle A | 0 | 842 | 20 | -0.2856 | 0.4984 | 0.2082 | 1 |
| Inferior Facial Angle B | 0 | 856 | 20 | -0.0599 | 0.5459 | 0.7922 | 1 |
| Inferior Facial Angle C | 0 | 837 | 20 | 0.21 | 0.5585 | 0.3564 | 1 |
| Inferior Facial Angle D | 0 | 850 | 20 | 0.4069 | 0.6593 | 0.0762 | 1 |
| Mentolabial Angle | 0 | 856 | 20 | -0.1464 | 0.5968 | 0.5204 | 1 |
| Philtral Depth | 0 | 877 | 20 | -0.124 | 0.5009 | 0.5843 | 1 |
| Philtral Depth | 1 | 876 | 20 | -0.0319 | 0.5538 | 0.8888 | 1 |
| Philtral Depth | 2 | 872 | 20 | -0.2079 | 0.414 | 0.3566 | 1 |
| Orbital Protrusion | 0 | 802 | 20 | -0.0893 | 0.543 | 0.6946 | 1 |
| Nasal Root Depth | 0 | 878 | 20 | -0.3203 | 0.3845 | 0.1549 | 1 |
| Bifrontal Asymmetry | 0 | 802 | 20 | 0.0524 | 0.3815 | 0.816 | 1 |
| Bizygomatic Asymmetry | 0 | 852 | 19 | -0.185 | 0.3501 | 0.4227 | 1 |
| Bizygomatic Asymmetry | 2 | 846 | 19 | -0.1392 | 0.3932 | 0.5471 | 1 |
| Cheekbone Asymmetry | 0 | 870 | 20 | -0.3724 | 0.7641 | 0.1073 | 1 |
| Cheekbone Asymmetry | 1 | 867 | 20 | -0.0471 | 0.7381 | 0.8382 | 1 |
| Nasal Ala Asymmetry | 0 | 870 | 20 | -0.3983 | 0.3739 | 0.0773 | 1 |
| Nasal Asymmetry | 0 | 870 | 20 | -0.3825 | 0.3873 | 0.0899 | 1 |
| Nasal Asymmetry | 1 | 874 | 20 | -0.2155 | 0.4386 | 0.34 | 1 |
| Subnasal Asymmetry | 0 | 873 | 20 | -0.3666 | 0.3218 | 0.103 | 1 |
| Subnasal Asymmetry | 1 | 877 | 20 | -0.1156 | 0.3406 | 0.6071 | 1 |
| Subnasal Asymmetry | 2 | 877 | 20 | 0.1301 | 0.5058 | 0.5663 | 1 |
| Nasal Root Asymmetry | 0 | 868 | 20 | -0.1531 | 0.3308 | 0.4963 | 1 |
| Nasal Root Asymmetry | 1 | 874 | 20 | 0.0227 | 0.5349 | 0.9201 | 1 |
| Nasal Root Asymmetry | 2 | 873 | 20 | 0.3059 | 0.3192 | 0.1737 | 1 |
| Nasal Root Asymmetry | 3 | 867 | 20 | -0.0778 | 0.2906 | 0.7293 | 1 |
| Philtral Asymmetry | 0 | 871 | 20 | -0.1806 | 0.5406 | 0.4268 | 1 |
| Philtral Asymmetry | 1 | 873 | 20 | -0.2459 | 0.5167 | 0.2787 | 1 |
| Philtral Asymmetry | 2 | 873 | 20 | -0.2498 | 0.4693 | 0.2698 | 1 |
| Philtral Asymmetry | 3 | 867 | 20 | 0.4946 | 0.4634 | 0.029 | 1 |
| Labial Fissure Asymmetry | 0 | 870 | 20 | -0.3674 | 0.4895 | 0.1051 | 1 |
| Labial Fissure Asymmetry | 1 | 876 | 20 | -0.5067 | 0.4335 | 0.025 | 1 |
| Labial Fissure Asymmetry | 3 | 869 | 20 | 0.2198 | 0.4612 | 0.3312 | 1 |
| Innercanthal Asymmetry | 0 | 867 | 20 | -0.5621 | 0.4489 | 0.0131 | 0.9406 |
| Innercanthal Asymmetry | 1 | 875 | 20 | -0.3141 | 0.4972 | 0.166 | 1 |
| Innercanthal Asymmetry | 2 | 875 | 20 | -0.0833 | 0.5537 | 0.7142 | 1 |
| Innercanthal Asymmetry | 3 | 864 | 20 | 0.4804 | 0.4865 | 0.0342 | 1 |
| Outercanthal Asymmetry | 0 | 870 | 20 | 0.0883 | 0.4454 | 0.6961 | 1 |
| Outercanthal Asymmetry | 1 | 873 | 20 | 0.0935 | 0.5848 | 0.6815 | 1 |
| Outercanthal Asymmetry | 2 | 870 | 20 | 0.1949 | 0.396 | 0.3876 | 1 |
| Outercanthal Asymmetry | 3 | 867 | 20 | 0.2425 | 0.3527 | 0.2813 | 1 |
| Mid-Infraorbital Asymmetry | 0 | 866 | 20 | 0.1669 | 0.7722 | 0.4704 | 1 |
| Mid-Infraorbital Asymmetry | 1 | 866 | 20 | -0.2778 | 0.5439 | 0.2218 | 1 |
| Mid-Infraorbital Asymmetry | 2 | 865 | 20 | 0.3724 | 0.3786 | 0.0986 | 1 |
| Mid-Infraorbital Asymmetry | 3 | 861 | 20 | 0.3412 | 0.4677 | 0.1317 | 1 |
| Mid-Supraorbital Asymmetry | 0 | 871 | 20 | 0.1483 | 0.5951 | 0.5156 | 1 |
| Mid-Supraorbital Asymmetry | 2 | 868 | 20 | 0.2434 | 0.4328 | 0.2813 | 1 |
| Temples Asymmetry | 1 | 797 | 20 | 0.1316 | 0.3181 | 0.5583 | 1 |

